# Financial Hardship and Social Assistance as Determinants of Mental Health and Food and Housing Insecurity During the COVID-19 Pandemic in the United States

**DOI:** 10.1101/2020.12.24.20248835

**Authors:** Daniel Kim

## Abstract

**Background:** While social assistance through the United States (U.S.) federal Coronavirus Aid, Relief, and Economic Security (CARES) Act provided expanded unemployment insurance benefits during the coronavirus disease 2019 (COVID-19) pandemic until the summer of 2020, it is unclear whether subsequent social assistance has been or will be sufficient to meet everyday spending needs and to curb the adverse health-related sequelae of financial hardship.

**Methods:** This study estimated recent trends in financial hardship among working-aged Americans with job-related income loss during the pandemic. It also used multivariable logistic regression and repeated cross-sectional individual-level U.S. Household Pulse Survey data on 91,222 working-aged adults between September and December 2020 to explore the associations of financial hardship with mental health outcomes and food and housing insecurity after accounting for receipt of social assistance.

**Results:** Experiencing somewhat of a financial hardship (vs no hardship) was linked to 3-7 times higher odds of anxiety and depressive symptoms and a likely eviction, and 11 times higher odds of food insufficiency. Experiencing considerable financial hardship (vs no hardship) predicted 5-7 fold higher odds of anxiety and depressive symptoms, 34-fold higher odds of a likely housing eviction, and 37-fold higher odds of food insufficiency (all *P* values <.001). Across outcomes, these relationships were stronger at each successively higher level of financial hardship (all *P* values for linear trend <.001), and more than offset any corresponding benefits from social assistance.

**Conclusions:** Even after accounting for receipt of social assistance, working-aged adults experiencing financial hardship had markedly greater odds of anxiety and depressive symptoms, food insufficiency, and an anticipated housing eviction. These findings point to the urgent need for direct and sustained cash relief well in excess of current levels of social assistance, and provide a critical baseline assessment for evaluating the impacts of federal public policy responses to economic hardships during the pandemic. It is essential that the U.S. Congress and the new Biden administration provide adequate and needs-based social policy relief measures in order to mitigate the pandemic’s adverse impacts on the physical, mental, and social well-being of millions of Americans.

## Introduction

The coronavirus disease 2019 (COVID-19) pandemic has led to the filing of over 70 million unemployment insurance (UI) claims in the United States (U.S.),^1^ and has been accompanied by heightened levels of food and housing insecurity.^2^ Through the federal Coronavirus Aid, Relief, and Economic Security (CARES) Act enacted in March 2020, expanded UI benefits consisted of a $600 weekly payment on top of state payments, 13 extra weeks of UI benefits, and broader UI eligibility guidelines.^3^ The federal bonus expired in late July 2020, and was followed by a six-week $300 weekly benefit in the majority of states through a federal lost wages assistance program.^4^ Other provisions along with a public health housing eviction moratorium implemented in September 2020 expired in late December 2020. A few days later, both the U.S. Congress and President Trump approved a $900 billion COVID-19 relief package that included a one-time direct payment of $600 to individuals (with a 2019 adjusted gross income of up to $75,000) and a federal UI bonus of $300 weekly for 11 weeks.^5^ In mid-January 2021, President-elect Biden unveiled an $1.9 trillion economic relief plan for his administration that proposed a one-time direct payment of $1,400 (on top of the $600 check) and a UI supplement of $400 weekly through the end of September 2021;^6^ this proposal has yet to be approved by Congress.

An analysis of data from the U.S. Census Bureau’s Household Pulse Survey (HPS) administered in June and July 2020 identified UI receipt as associated with better mental health and lower health-related social needs among working-aged adults.^7^ However, since the federal UI bonus subsequently lapsed and was followed by a subsidy considerably smaller in both amount and duration, it is unclear whether social assistance has been or will be sufficient to meet everyday spending needs and to curb adverse sequelae of financial hardship among those experiencing employment-related income loss. This study was undertaken to estimate recent levels of and changes in financial hardship among working-aged Americans with job-related income loss during the most recent months of the pandemic, and to explore the associations of financial hardship with mental health outcomes and food and housing insecurity after accounting for receipt of social assistance.

## Methods

### Study population

Repeated cross-sectional individual-level data were pooled from nationally-representative HPS surveys administered from September 2—December 21, 2020.^8^ The study population was adults age 18-64 years reporting a loss of household employment income since the beginning of the pandemic (March 13, 2020). The most recently available aggregate data were also drawn from the HPS public-use survey data tables (December 9—December 21). The HPS used the U.S. Census Bureau’s Master Address File as the source of sampled housing units (HUs). The sampling frame was a systematic sample of all eligible HUs. The HPS was conducted online by Qualtrics as the data collection platform. Across data collection periods, survey response rates ranged from 5.3% to 10.6%.^9^

### Predictors

Financial hardship was defined as household difficulty (not at all/a little/somewhat/very difficult) to pay for usual household expenses including food, rent/mortgage, and loans within the previous week. UI receipt was taken as household receipt of UI benefits since the pandemic began. Supplemental Nutritional Assistance Program (SNAP) receipt was defined as household participation in SNAP and using SNAP benefits to meet spending needs within the previous week.^9^

### Outcomes

Outcomes consisted of the frequency of anxiety and depressive symptoms (using the 2-item Generalized Anxiety Disorder-2 and Patient Health Questionnaire-2, respectively, for scores of 3-6 vs 0-2 to screen for anxiety and depressive disorders, with acceptable sensitivity and specificity levels^10^), the level of food insufficiency (often or sometimes not enough to eat vs enough to eat), and, among housing renters, the likelihood of being evicted within the next 2 months (very/extremely likely vs somewhat/not at all likely).^9^

### Statistical analysis

Multivariable logistic regression models were fit to estimate adjusted odds ratios from generalized estimating equations that accounted for repeated measures within individuals and person-level survey weights and provided robust standard errors. Log-Poisson regression was not employed due to potential bias when estimating relative risks for binary outcomes.^11^ All models were adjusted for individual age, age squared, gender, race/ethnicity, marital status, education, 2019 household income, other federal stimulus assistance, household size, presence of children in the household, general health status, state, and survey period. The model for current food insufficiency was also controlled for pre-pandemic food insufficiency. Multiple imputation analysis (using 25 multiply imputed datasets) and complete case analysis were used to handle missing data for predictor and outcome variables, respectively. All analyses were conducted using SAS version 9.4 (SAS Institute Inc., Cary, NC).

## Results

*Figure 1* shows the estimated numbers and percentages of U.S. working-aged adults with employment income loss, receipt/denial of UI benefits, mental health-related symptoms, food insufficiency, and financial hardship in the latest wave of the HPS (December 9—December 21). Over half (55.5%) of working-aged adults (representing more than 108 million adults) experienced job-related income loss in their households since the start of the pandemic. Of those who lost employment income, less than half (46.6%) applied for UI benefits, and of those who applied, 76.6% received UI. More than one-third (37.8%) and one-quarter (28.5%) reported a higher frequency of feeling anxious or depressed, respectively. 16.0% (corresponding to 27 million adults) experienced food insufficiency. Over half (51.7%) of renters (representative of 4.8 million adults) indicated a high likelihood of being evicted. 42.0% (corresponding to 79 million adults) reported a higher level of financial hardship. All proportions were at least 5 percentage points higher among those with employment income loss *(Figure 1)*.

**Figure 1.**
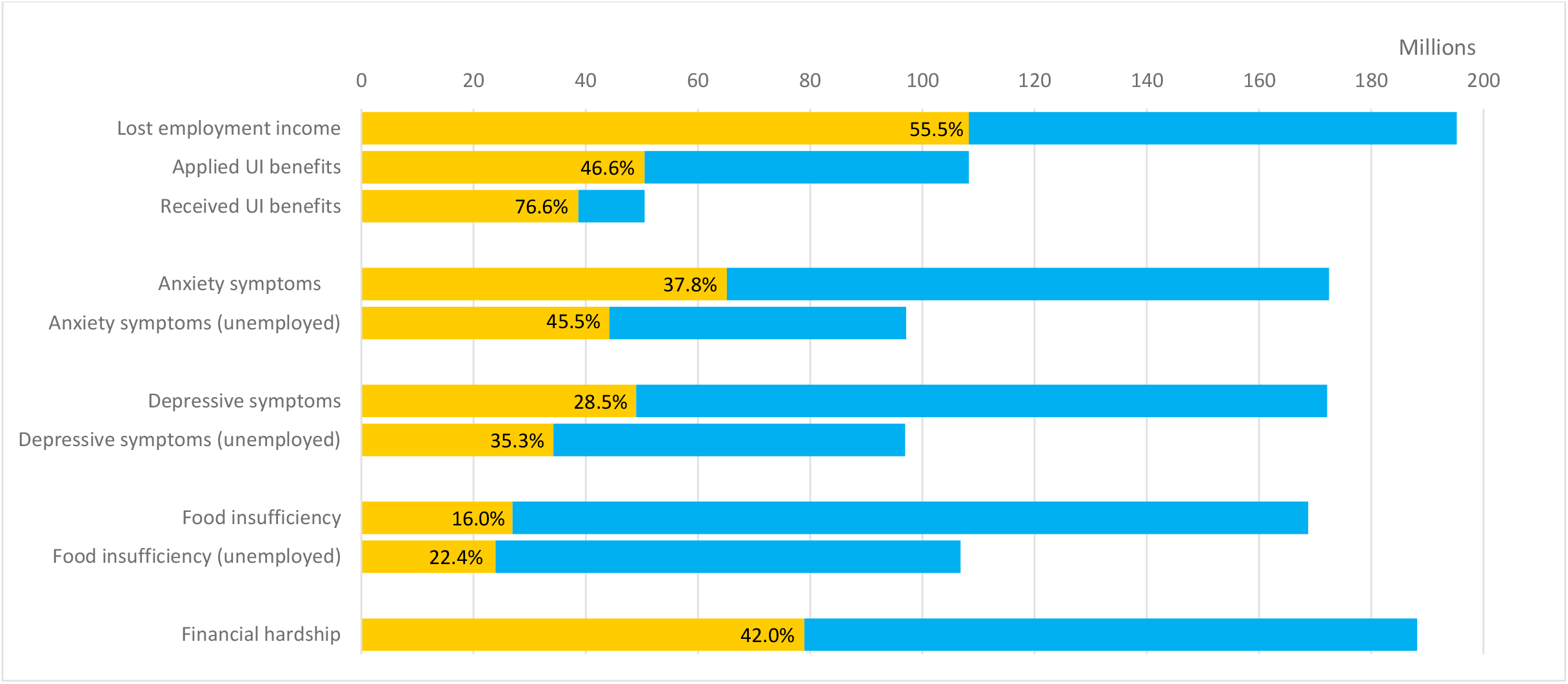
Estimated Numbers and Percentages of Working-Aged Adults with Household Employment Income Loss, Receipt/Denial of UI Benefits, Mental Health-Related Symptoms, Food Insufficiency, and Financial Hardship During the COVID-19 Pandemic, U.S. Census Bureau Household Pulse Survey, December 2020 ^a^. Abbreviations: COVID-19, coronavirus disease 2019; UI, unemployment insurance. ^a^ Aggregate data were drawn from the latest U.S. Census Bureau Household Pulse Survey public-use data tables for surveys administered between December 9, 2020 and December 21, 2020. All estimates are for adults age 18-64 years except anxiety and depressive symptoms, for which available estimates are for adults age 18-69 years. The yellow bars indicate the number of adults (in millions), and are accompanied by percentage estimates reflecting the number of adults as a percentage of the total number of adults in the denominator (e.g., number responding someone in household lost employment income, number who applied for UI benefits, number who responded to the survey item). Anxiety symptoms were measured by a survey item that inquired about the frequency of feeling nervous, anxious, or on edge over the previous week. Depressive symptoms were measured by a survey item that inquired about the frequency of feeling down, depressed, or hopeless over the previous week. Financial hardship corresponded to it being somewhat to very difficult (vs not at all or a little difficult) to pay for usual expenses over the previous week.

*Figure 2* depicts the estimated national percentages of working-aged adults experiencing household financial hardship (somewhat to very difficult to pay for usual household expenses) by survey period between September 2, 2020 and December 21, 2020. In September 2020, this percentage declined slightly from 36.3% to 35.5%, but then rose progressively during each successive survey period to 42.0% by December 2020. The largest absolute percentage change (+1.4 percentage points) occurred between the two most recent survey periods, suggesting the possibility of an escalating trend (*Figure 2*).

**Figure 2.**
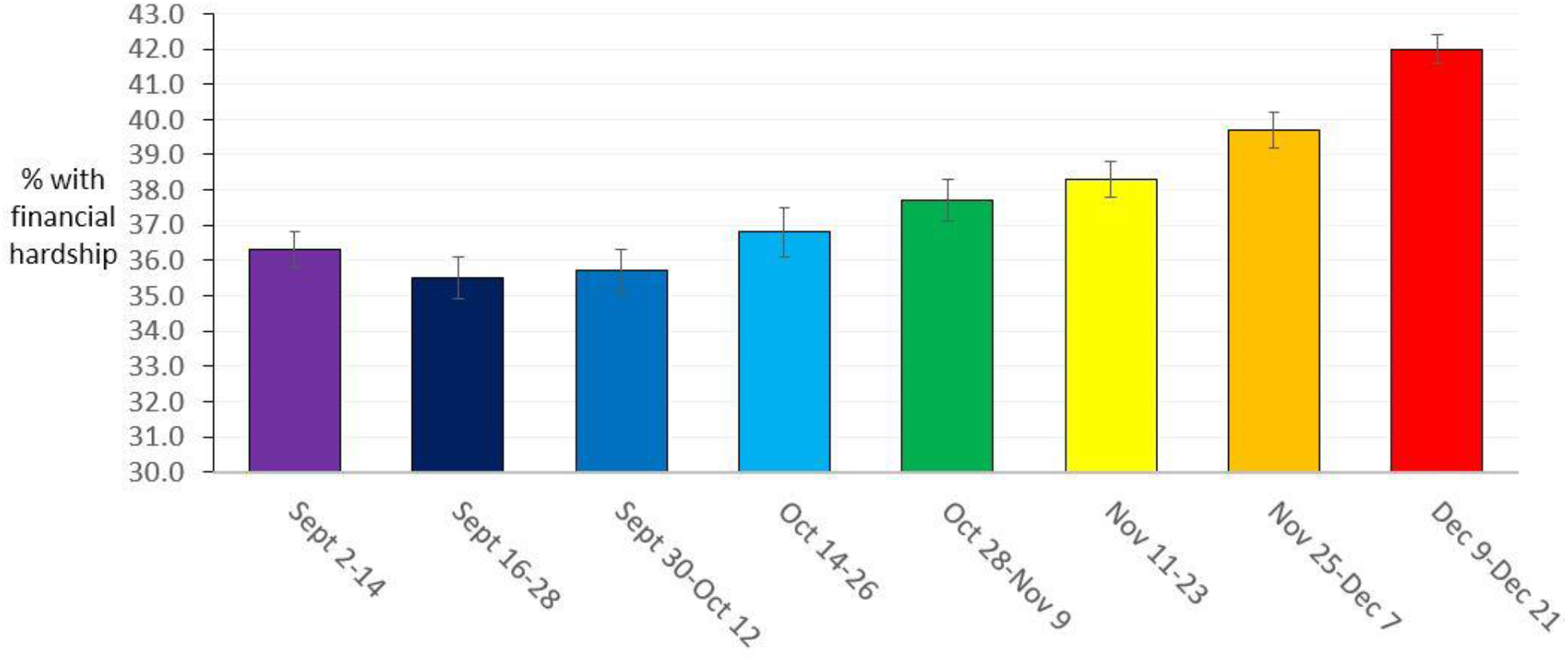
Estimated Percentages of Working-Aged Adults Experiencing Household Financial Hardship During the COVID-19 Pandemic by Survey Week, U.S. Census Bureau Household Survey, September-December 2020.^a^. ^a^ Aggregate data were drawn from the U.S. Census Bureau Household Pulse Survey public-use data tables for surveys administered between September 2, 2020 and December 21, 2020. All estimates are for adults age 18-64 years. Financial hardship corresponded to it being somewhat to very difficult (vs not at all or a little difficult) to pay for usual expenses over the previous week.

*Table 1* displays the main results from the multivariable-adjusted models. In models using data on up to 91,222 working-aged adults (representative of 43 million individuals) with income disruption, experiencing somewhat of a financial hardship (vs no hardship) was linked to 3-7 times higher odds of anxiety and depressive symptoms and a likely eviction, and 11 times higher odds of food insufficiency. Experiencing considerable financial hardship (vs no hardship) predicted 5-7 fold higher odds of anxiety and depressive symptoms, 34-fold higher odds of a likely eviction, and 37-fold higher odds of food insufficiency (*Table 1*; all *P* <.001; all *P* for linear trend <.001). In the same models, UI receipt (vs no receipt) was associated with 9-12% lower odds of depressive symptoms (*P* = 0.03), anxiety symptoms (*P* = 0.005), and food insufficiency (*P* = 0.10), and 35% lower odds of an expected eviction (*P* <.001). SNAP receipt was linked to 10% lower odds of food insufficiency (*P* = 0.19).

**Table 1.** **Financial Hardship and Social Assistance as Predictors of Mental Health, Food Insufficiency, and Likely Housing Eviction Among Those with Job-Related Income Loss During the COVID-19 Pandemic, U.S. Census Bureau Household Pulse Survey, September-December 2020^a^**

|  | Outcome |  |  |  |  |  |  |  |
| --- | --- | --- | --- | --- | --- | --- | --- | --- |
|  | Anxiety symptoms |  | Depressive symptoms |  | Food insufficiency |  | Likely housing eviction<sup>c</sup> |  |
| Predictor | Odds ratio (95% CI)<sup>b</sup> | P value | Odds ratio (95% CI)<sup>b</sup> | P value | Odds ratio (95% CI)<sup>b</sup> | P value | Odds ratio (95% CI)<sup>b</sup> | P value |
| Difficulty with Paying Expenses<sup>d</sup> |  |  |  |  |  |  |  |  |
| - *A little difficult* | 1.94 (1.76-2.14) | <.001 | 1.71 (1.52-1.92) | <.001 | 4.04 (2.73-5.97) | <.001 | 1.98 (0.71-5.52) | .19 |
| - *Somewhat Difficult* | 3.03 (2.75-3.35) | <.001 | 2.70 (2.41-3.03) | <.001 | 10.56 (7.29-15.30) | <.001 | 6.54 (2.48-17.29) | <.001 |
| - *Very difficult* | 6.64 (5.99-7.35) | <.001 | 5.43 (4.83-6.10) | <.001 | 36.94 (25.58-53.35) | <.001 | 33.89 (13.08-87.83) | <.001 |
| **P for trend** | - | <.001 | - | <.001 | - | <.001 | - | <.001 |
| Receipt of UI Benefits<sup>e</sup> | 0.88 (0.81-0.96) | .005 | 0.91 (0.83-0.99) | .03 | 0.90 (0.80-1.02) | .10 | 0.65 (0.55-0.77) | <.001 |
| Receipt of SNAP Benefits<sup>e</sup> | - | - | - | - | 0.90 (0.78-1.05) | .19 | - | - |

Abbreviations: CI, confidence interval; COVID-19, coronavirus disease 2019; SNAP, Supplemental Nutrition Assistance Program; UI, unemployment insurance.
<sup>a</sup> All models were adjusted for age, age<sup>2</sup>, gender, race/ethnicity, marital status, educational attainment, 2019 household income, use of federal stimulus assistance, household size, presence of children in household, overall health status, state of residence, and survey period. The model for current food insufficiency was also adjusted for food insufficiency prior to March 13, 2020. Participants were surveyed in Phases 2 and 3 of the U.S. Census Bureau Household Pulse Survey (Phase 2 waves: September 2—14, September 16—28, September 30—October 12, October 14—26; Phase 3 waves: October 28—November 9, 2020, November 11—November 23, 2020, November 25—December 7, 2020, December 9—December 21, 2020).
<sup>b</sup> Odds ratio point estimates, 95% confidence intervals, and *P* values were derived from logistic regression models fit using generalized estimating equations that incorporated person weights, repeated measures, and robust standard errors. Missing data were handled using 25 multiple imputation data sets.
<sup>c</sup> This analysis was restricted to housing renters.
<sup>d</sup> The reference category was those reporting it was not at all difficult to pay for usual expenses over the previous week.
<sup>e</sup> The reference category was those reporting no household receipt of corresponding UI or SNAP benefits since March 13, 2020.

## Discussion

This large U.S. nationally-representative study reveals high levels of financial hardship among working-aged Americans, that progressively worsened from September to December 2020. Even after accounting for social assistance receipt and pre-pandemic socioeconomic position, working-aged adults experiencing financial hardship had markedly greater odds of anxiety or depressive symptoms, food insufficiency, and an anticipated housing eviction. Across outcomes, these relationships were stronger at each successively higher level of financial hardship, and more than offset any corresponding benefits from UI or SNAP. To the author’s knowledge, this represents the first study to explore the potential impacts of financial hardship with mental health and food and housing insecurity during the COVID-19 pandemic in the United States. The associations with mental health outcomes are in keeping with previous linkages of sudden income loss to depression and anxiety in labor market surveys conducted in six European countries in March and April 2020.^12^

Strengths of the study include its use of large nationally-representative survey data, as well as data from repeated survey waves that enabled the examination of trends in financial hardship over time. Models also controlled for baseline demographic and socioeconomic factors and state fixed effects to reduce confounding. Finally, the modeling of multiple categories for the financial hardship measure permitted a confirmation of the presence of dose-response relationships.

Nonetheless, there are limitations to this study. Because of the study’s cross-sectional and observational design, bias due to reverse causation or confounding cannot be entirely ruled out. Moreover, we were unable to account for the received monetary amounts of UI and SNAP benefits. While sampling weights accounted for non-response, the low survey response rates could have led to selection bias. Last, all measures were based on self-report, although the adjustment for general health status in all models should have attenuated the degree of same-source bias.

There is evidence to support that during the period that the CARES Act was in effect during the initial phase of the pandemic, among adults whose families lost work or work-related income due to the pandemic, the level of social assistance including the $600 weekly federal bonus for UI recipients was adequate to meet financial needs.^13^ UI receipt was linked to a 3-percentage point reduction in the share reporting food insecurity, a 3.7 percentage-point reduction in problems paying utility bills, and reductions of 8.6 to 15.1 percentage points in the share worrying about meeting basic needs.^13^

Overall, the findings from the present study suggest the importance of financial hardship as a fundamental cause^14^ and social determinant of mental health and social needs during the COVID-19 pandemic, and point to the urgent need for direct and sustained cash relief well in excess of current levels of social assistance, as well as the imperative of extending housing renter eviction protections. The current study further offers a critical baseline assessment for evaluating impacts of the U.S. federal government’s public policy responses in subsequent months and years of the pandemic. Notably, this study also helps to heed recent calls for a more “consequential” epidemiology, whereby epidemiologic research serves to more directly inform contemporary social policies to improve population health, including in response to emerging public health threats and crises including pandemics.^15^

Based on the above prior evidence that economic hardship and food insecurity declined among UI recipients after enacting the CARES Act, and the present study’s evidence that a substantial and growing share of working-aged adults has experienced household financial hardship since federal UI supplements lapsed by September 2020, in order to alleviate financial hardship and its adverse sequelae, a logical recommendation is that any future federal UI bonus match or even exceed the UI bonus level of $600 weekly as well as a one-time direct payment of $1,200 previously provided through the CARES Act.^3^ The COVID-19 relief bill approved by the U.S. Congress and President Trump limited the federal UI supplement to $300 weekly and a one-time stimulus check of $600.^5^

The COVID-19 economic relief plan proposed in January 2021 by the new Biden administration would bring the recent direct one-time payments to individuals to a total of $2,000, and UI supplements to $400 weekly through the end of September 2021.^6^ Even if these elements of the Biden administration’s relief plan are approved by Congress, the relative $200 weekly shortfall in the UI supplement compared to the CARES Act would most likely signify that households experiencing income loss during the pandemic will not have their financial needs sufficiently met. Based on the strong associations demonstrated between financial hardship and physical and mental health outcomes in the present study, such a shortfall would likely carry substantial negative public health consequences. Over the ensuing months, and possibly years of the pandemic, it is essential that the U.S. Congress and the Biden administration provide expanded and adequate needs-based social policy relief measures in order to mitigate the pandemic’s short- and long-term adverse impacts on the physical, mental, and social well-being of tens of millions of Americans.

## Data Availability

All data are publicly available.

## Notes

### Competing Interest Statement

The authors have declared no competing interest.

### Funding Statement

No external funding was received for this work.

### Author Declarations

Because all data lacked identifying information and were publicly available through the U.S. Census Bureau, this study was deemed exempt by the Human Subject Research Protection Committee at Northeastern University.

### Summary of Updates

The analyses were updated to included additional periods of survey data from the U.S. Household Pulse Survey.

